# Global, Regional, and National Survey on Burden and Quality of Care Index (QCI) of Orofacial Clefts: Global Burden of Disease Systematic Analysis 1990–2019

**DOI:** 10.1101/2024.03.24.24304795

**Authors:** Ahmad Sofi-Mahmudi, Erfan Shamsoddin, Sahar Khademioore, Yeganeh Khazaei, Marcos Roberto Tovani-Palone

**Affiliations:** National Pain Centre, Department of Anesthesia, McMaster University, Hamilton, ON, Canada; Department of Health Research Methods, Evidence and Impact, McMaster University, Hamilton, ON, Canada; Seqiz Health Network, Kurdistan University of Medical Sciences, Seqiz, Kurdistan; Cochrane Iran Associate Centre, National Institute for Medical Research Development (NIMAD), Tehran, Iran; Department of Statistics, Statistical Consultation Unit, StaBLab, LMU Munich, Akademiestr. 1, 80799, Munich, Germany; Ribeirão Preto Medical School, University of São Paulo, São Paulo, 14049-900, Brazil

**Keywords:** Cleft palate, cleft lip, global burden of disease, quality of health care

## Abstract

**Background:** Orofacial clefts are the most common craniofacial anomalies that include a variety of conditions affecting the lips and oral cavity. They remain a significant global public health challenge. Despite this, the quality of care for orofacial clefts has not been investigated in global and country level.

**Objective:** We aimed to measure the quality of care index (QCI) for orofacial clefts worldwide.

**Methods:** We used the 2019 Global Burden of Disease data to create a multifactorial index (QCI) to assess orofacial clefts globally and nationally. By utilizing data on incidence, prevalence, years of life lost, and years lived with disability, we defined four ratios aimed at indirectly reflecting the quality of healthcare. Subsequently, we conducted a principal component analysis to identify the most critical variables that could account for the observed variability. The outcome of this analysis was defined as the QCI for orofacial clefts. Following this, we tracked the QCI trends among males and females worldwide, across various regions and countries, considering factors such as the socio-demographic index and World Bank classifications.

**Results:** Globally, the QCI for orofacial clefts exhibited a consistent upward trend from 1990 to 2019 (66.4 to 90.2) overall and for females (82.9 to 94.3) and males (72.8 to 93.6). In the year 2019, the top five countries with the highest QCI scores were as follows: Norway (QCI=99.9), Ireland (99.4), France (99.4), Germany (99.3), the Netherlands (99.3), and Malta (99.3). Conversely, the five countries with the lowest QCI scores on a global scale in 2019 were Somalia (59.1), Niger (67.6), Burkina Faso (72.6), Ethiopia (73.0), and Mali (74.4). Gender difference showed a converging trend from 1990 to 2019 (optimize gender disparity ratio (GDR): 123 vs. 163 countries), and the GDR showed a move toward optimization (between 0.95 and 1.05) in the better and worse parts of the world.

**Conclusion:** Despite the positive results regarding the QCI for orofacial clefts worldwide, some countries showed a slight negative trend.

## Introduction

Non-syndromic orofacial clefts, which include cleft lip, cleft lip and palate, and cleft palate alone, comprise a range of disorders affecting the lips and oral cavity, the causes of which remain largely unknown (1). Considering that several distinct etiologies have been suggested for orofacial clefts, they can appear in multiple forms: cleft lip alone without cleft palate, cleft lip with or without cleft palate, cleft lip with cleft palate, posterior cleft palate without cleft lip. Orofacial clefts can also be categorized as syndromic and non-syndromic, or familial and nonfamilial (simplex clefts) (2, 3).

Orofacial clefts are estimated to occur in approximately 1 out of every 700 births and represent an enormous public health burden worldwide (4–7). Effects on speech, hearing, appearance, and cognition can lead to long-lasting adverse outcomes for health and social integration (1). Affected children need multidisciplinary care from birth until adulthood and have higher morbidity and mortality throughout life than unaffected individuals (8, 9). Findings of studies have shown an increased frequency of structural brain abnormalities (10) and that many children and their families are affected psychologically to some extent (11). Although rehabilitation is possible with good quality care, orofacial clefts inevitably pose a burden to the individual, the family, and society, with substantial expenditure on health and related services.

Care for children born with these defects generally includes many disciplines, namely nursing, plastic surgery, maxillofacial surgery, otolaryngology, speech therapy, audiology, counselling, psychology, genetics, orthodontics, and dentistry. Nevertheless, it forms only a part of every area’s clinical load, meaning that care has tended to be fragmented. This fragmentation of care has led to substantial variations in management, which continue to cause controversy. Furthermore, in both developing and developed countries, care standards for patients with cleft lip, cleft lip and palate, or cleft palate alone remain a cause for concern (12, 13).

The Global Burden of Disease (GBD) 2019 study estimates that the global disability-adjusted life years (DALYs) rate for orofacial clefts has decreased by more than 55% since 1990. In order to assess and quantify this progress achieved in addressing the global burden of orofacial clefts, the introduction of a comprehensive metric like the Quality of Care Index (QCI) could play a pivotal role. The QCI offers a structured framework to evaluate the effectiveness and adequacy of healthcare interventions, considering factors such as access to medical services, surgical outcomes, patient satisfaction, and follow-up care. By incorporating the QCI into the evaluation framework, policymakers and healthcare professionals can gain valuable insights into the quality of care provided to individuals with orofacial clefts over time. The concept of a QCI also finds support in various healthcare studies, where it has proven to be a useful tool for guiding healthcare policy decisions and resource allocation.

In this study, we aim to estimate the quality of care for orofacial clefts between 1990 and 2019 by utilizing an original methodology. This innovative approach could address the existing gap in data availability and enable a more nuanced understanding of the disparities in orofacial cleft care across different countries and regions, and contribute to the enhancement of evidence-based policymaking for orofacial cleft management.

## Methods

The protocol of this descriptive study was published beforehand on the Open Science Framework (OSF) website (https://osf.io/94wr7). All code and data related to the study were shared via its OSF repository (https://osf.io/r94ch/) and GitHub (https://github.com/choxos/qci-cleft) at the time of submission of the manuscript. To ensure transparency and facilitate the reproducibility of our analyses, a PDF document containing the codes and corresponding outputs is provided in Appendix 1.

The source of data for this study was the GBD 2019 study, accessible to the public at http://ghdx.healthdata.org/gbd-2019. GBD 2019 offers a standardized approach to estimating incidence, prevalence, deaths, years of life lost (YLLs) due to premature mortality, years lived with disability (YLDs) also referred to as years lived in less-than-ideal health, and DALYs categorized by cause, age groups, sex, year, and location. A comprehensive explanation of the GBD study, including inputs, analytical processes, outcomes, and cause-specific methodologies, can be found elsewhere (14).

Identification of orofacial clefts in this analysis adhered to the International Statistical Classification of Diseases and Related Health Problems, 10th revision (ICD-10) as Q37.

Four indices related to the quality of care are described as follows:

1. 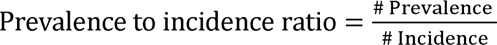
2. 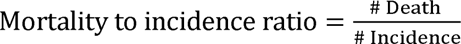
3. 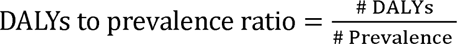
4. 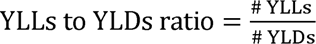

It is easy to understand the direction of these indices each one;. for example, if the ratio of how many people have the condition (prevalence) to how many new cases appear (incidence) is low, it could mean that care and prevention are betteror even could be a sign of decreased lifespan among the orofacial clefts patients. Lower mortality-to-incidence ratio shows better effectiveness of the provided care (if any). When the ratio of DALYs to prevalence is high, it means that the disease has a high burden (due to mortality or morbidity) in the country. The ratio of YLLs to YLDs indicates the mortality impact of the disease, with elevated figures reflecting a more compromised survival status for patients with orofacial clefts.To unify these indices, we utilized principal component analysis (PCA), as a multivariable analytical procedure. This technique extracts linear combinations of variables as orthogonal or uncorrelated components (15). The first-ranked component of the PCA, which is a linear combination of all variables, captures the majority of the variables’ information and was considered as the Quality-of-Care Index (QCI). The scores for these components were computed on a scale from 0 to 100, where higher values signify a better status.

We evaluated the distribution by employing the Socio-demographic Index (SDI), a concise metric reflecting a region’s developmental status. This assessment relies on the rankings of average per capita incomes, educational attainment levels, and fertility rates among all areas featured in the GBD study. Additionally, we incorporated World Bank classifications pertinent to global regions and individual countries. Furthermore, quintiles were employed on a yearly basis to depict the QCI scores of countries. To identify gender inequality within each country,, we used the gender disparity ratio (GDR), which simply is the male-to-female ratio of QCI. Five quintiles were defined as follows: 0 to 0.5, 0.5 to 0.95, 0.95 to 1.05, 1.05 to 1.5, and more than 1.5. Our preference was 0.95 to 1.05 quintile as the optimal GDR category. We employed a six-sigma test to find countries with a very high or very low QCI compared to others. The six-sigma approach calculates the mean and standard deviation of the index and specifies values out of the range of (μ-3σ, μ+3σ) as the outliers. The outliers, which pertain to countries with extreme values, can be interpreted in two ways: they might indicate underperformance in a particular context, such as during disease outbreaks, or they might reflect regions where the occurrence of a condition is unnaturally high or low. Full details of the analytical methods used in this study can be found elsewhere (16). From here onwards, except for the absolute values, all the DALY rates and QCIs reported in this paper represent age-standardized figures.

### Validation

Using a mixed-effect regression model, we considered QCI as a dependent variable. The independent variables were as follows: inpatient and outpatient healthcare utilization, orofacial clefts death, prevalence, and attributed death to all risk factors (17). Considering countries as a random effect, the Pearson’s correlation coefficient between the predicted QCI and healthcare access and quality of care index (HAQI) —an index to appraise the accessibility of care—was calculated to be 0.6 (18). All the analyses were performed using R 4.2.2. Detailed information on our mathematical model’s steps and the statistical protocol are available elsewhere (19).

## Results

### Burden

Globally, the age-standardized DALYs rate for the orofacial cleft was 19.63 (95% UI 12.85– 27.44 per 100,000) in 1990. DALYs rate for males was higher than females (21.04 (95% UI: 10.45–35.28) compared with 18.14 (95% UI: 12.12–35.68) per 100,000) in 1990. These numbers have decreased through the years, reaching an average overall DALYs rate of 7.51 (95% UI: 5.10–11.57), 7.58 (95% UI: 5.03–13.44) for males, and 7.45 (95% UI: 4.84–12.73) for females in 2019. The sharpest decrease in DALYs rate happened in the WB upper-middle-income (−83.2%) and high-middle SDI (−82.6%) countries; however, all regions experienced decreasing DALYs since 1990.

### Quality of care index and gender inequity

Global QCI for orofacial clefts gradually increased from 1990 to 2019 (from 66.3 to 90.2). In 1990, QCI was higher in females than males (82.9 vs 72.7). The quality of care index for both males and females globally increased throughout these years, reaching 93.6 for males and 94.3 for females, which means the gap between sexes narrowed down (Figure 1).

**Figure 1.**
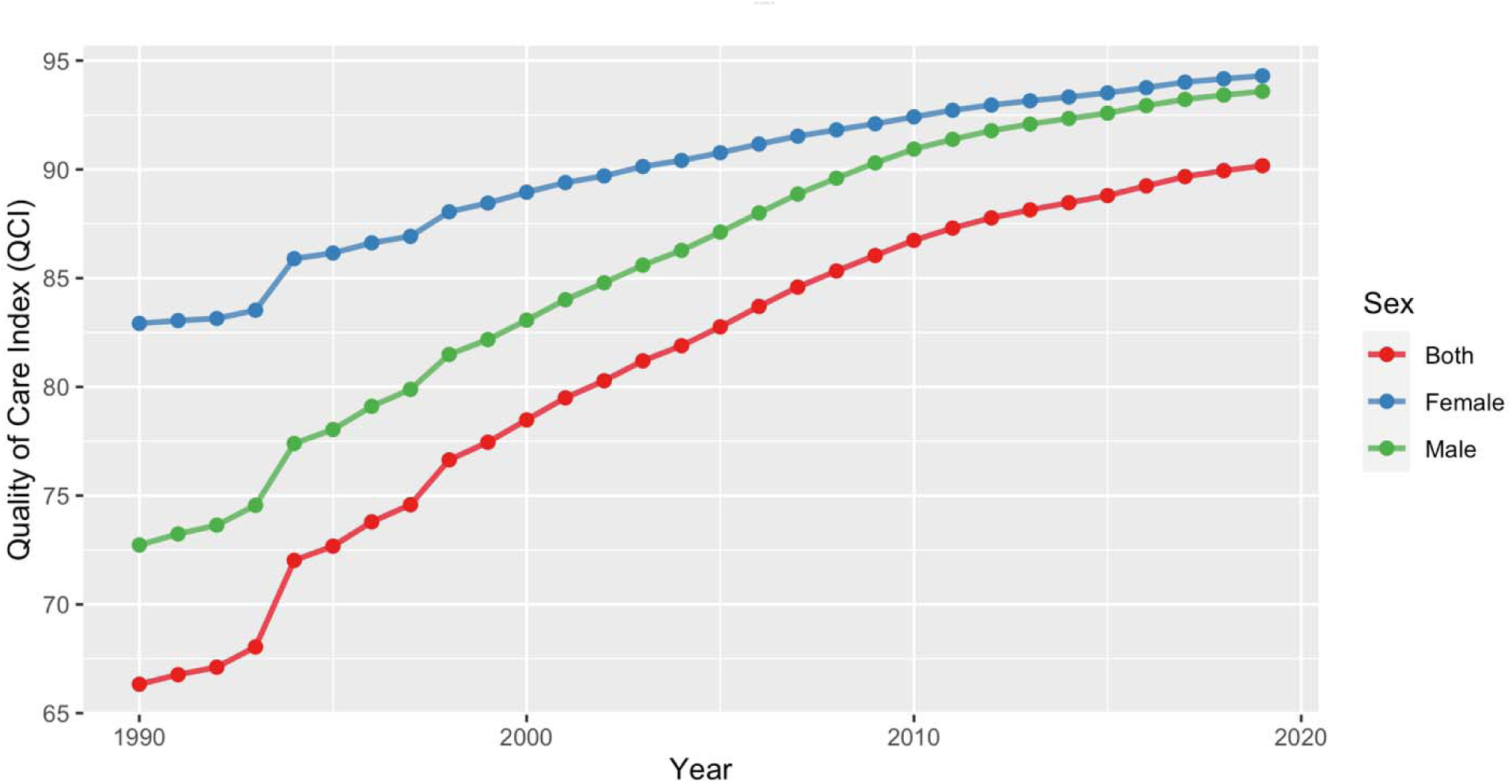
Time pattern of the age-standardized QCI (%) for orofacial clefts by gender between 1990 and 2019. QCI: Quality of care index.

When comparing the GDR among different countries, in 1990, in most African and Central Asian countries and some countries in South America, males received better care than females. However, in 2017, the majority of countries in the world provided equal care for both sexes. The global distribution of GDR in men and women in 1990 and 2017 is illustrated in Figure 2.

**Figure 2.**
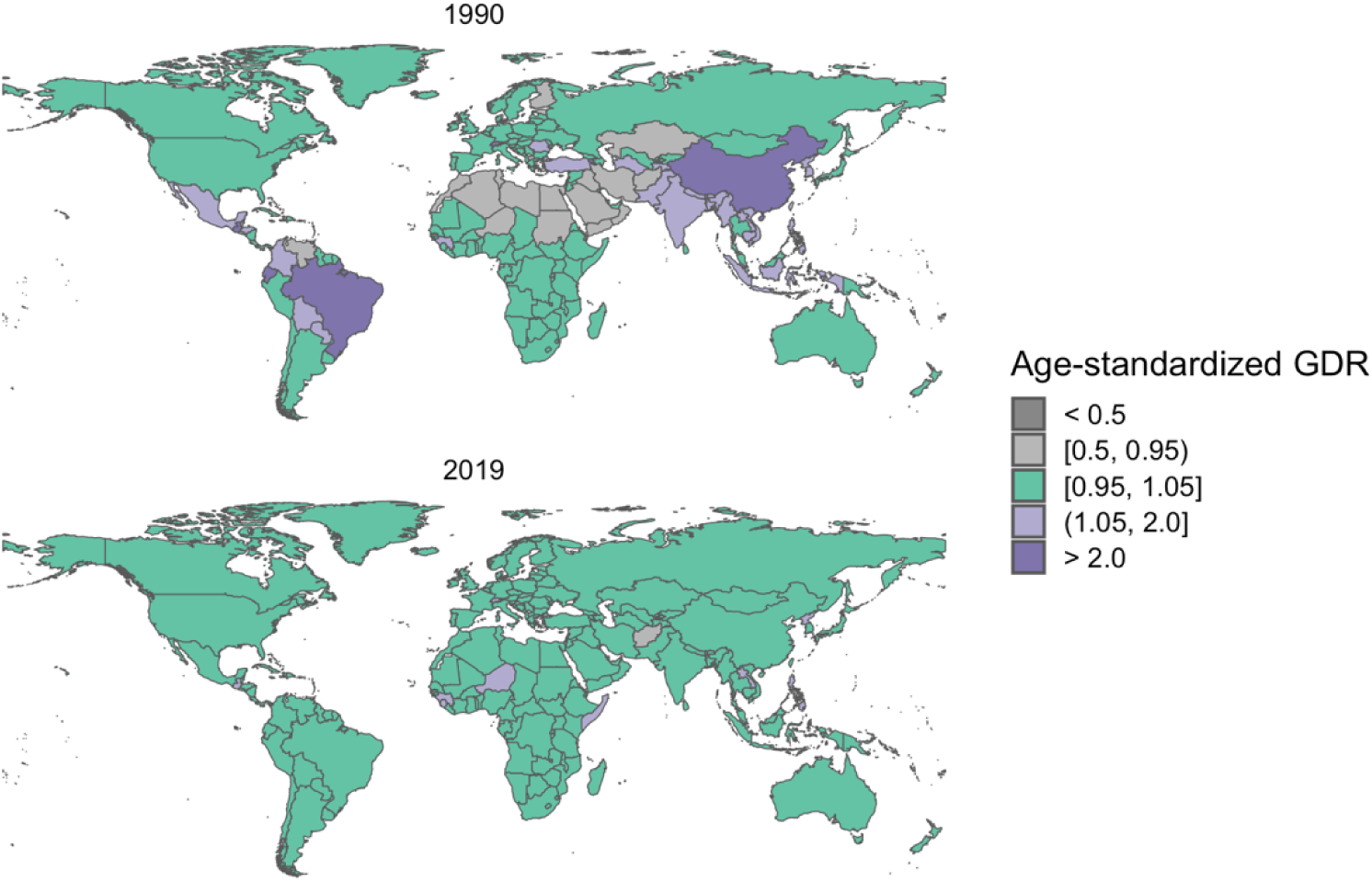
Geographical distribution of GDR for lip and oral cavity cancer. A: Age-standardised gender disparity ratio in men and women in 1990, B: Age-standardised gender disparity ratio in men and women in 2019. GDR: Gender disparity ratio.

### Comparison between countries

Between 1990 and 2019, the QCI for orofacial clefts increased in all the regions (either categorized SDI or World Bank) and countries, except for eight countries (4.0%). All these eight had a less than 1% change except for Zimbabwe (QCI change=–4.8). In 1990, Norway had the highest QCI (100.0), while Brazil had the lowest QCI (0.0). In 2019, Norway (99.9) and Somalia (59.1) had the highest and lowest QCI, respectively (Figure 3).

**Figure 3.**
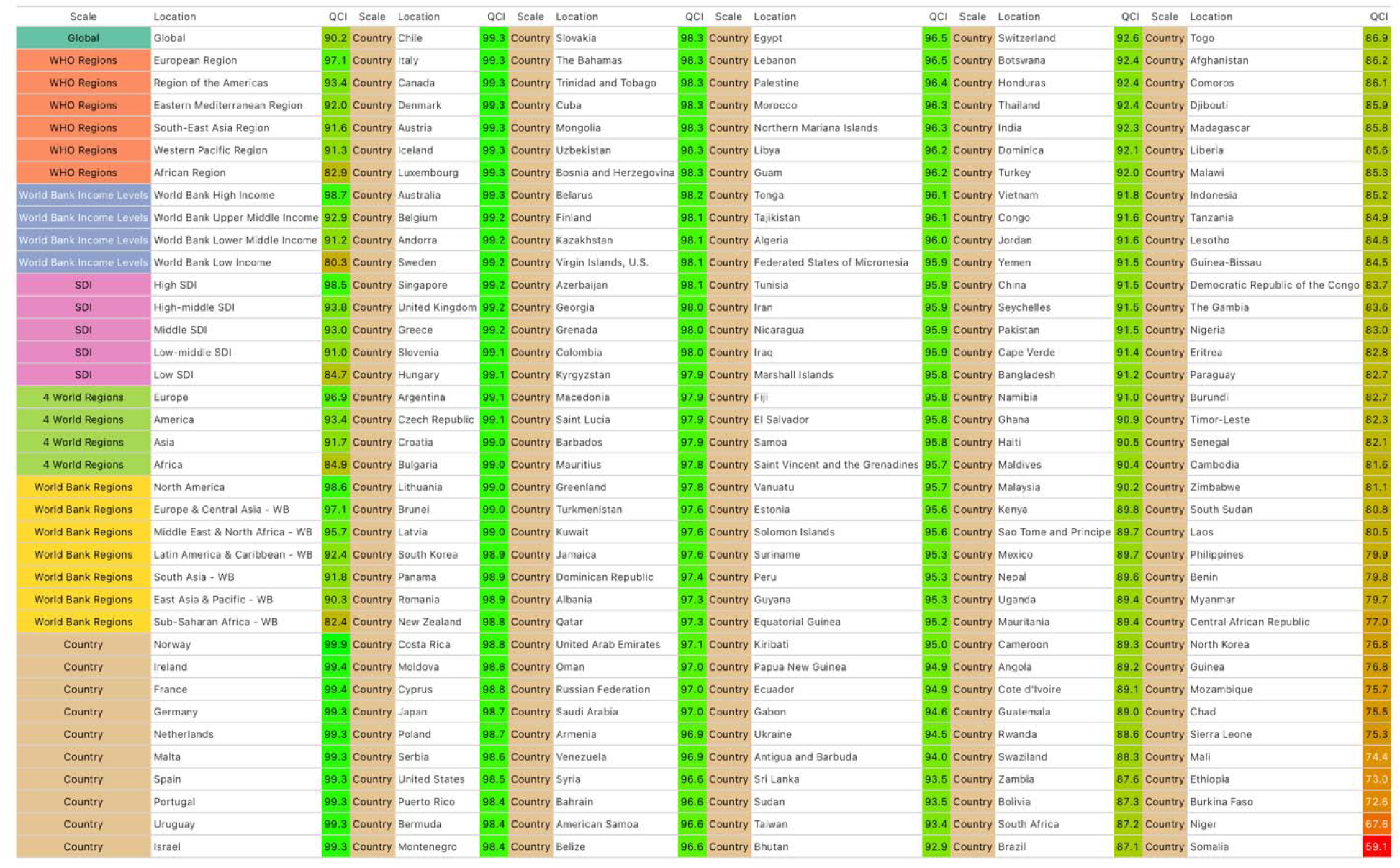
Global regions and countries listed in descending order based on their QCIs.

In 2019, the five countries with the least amount of QCIs were all African, while European, North American, and Oceanian countries had the highest QCIs (Figure 3). Globally, Norway ranked first regarding QCI (99.9), followed by Ireland (99.4), France (99.4), Germany (99.3), and the Netherlands (99.3).

Shifting to the other side of the QCI spectrum, Somalia (59.1), Niger (67.6), Burkina Faso (72.6), Ethiopia (73.0), and Mali (74.4) were the countries with the lowest figures. Figure 3 lists all the countries based on their QCIs.

Based on the World Bank Income Levels classification, high-income countries had the highest QCI in 2019 (=98.7), and low-income countries had the lowest QCI in the same year (=80.3). Based on the SDI classification, high SDI countries had the highest QCI in 2019 (=98.5), and low SDI countries had the lowest QCI in the same year (=84.7). However, World Bank upper-middle-income countries (54.6) and high-middle SDI countries (50.9) had the highest QCI increase rate (Table 1). In 1990, QCI in African, Asian, and South American countries was in the lowest quintile, while European and Oceanian countries had the highest QCI. In 2019, QCI in African countries was in the lowest quintile, while QCI in European, Oceanian, and North American countries was in the highest quintile (Figure 4).

**Figure 4.**
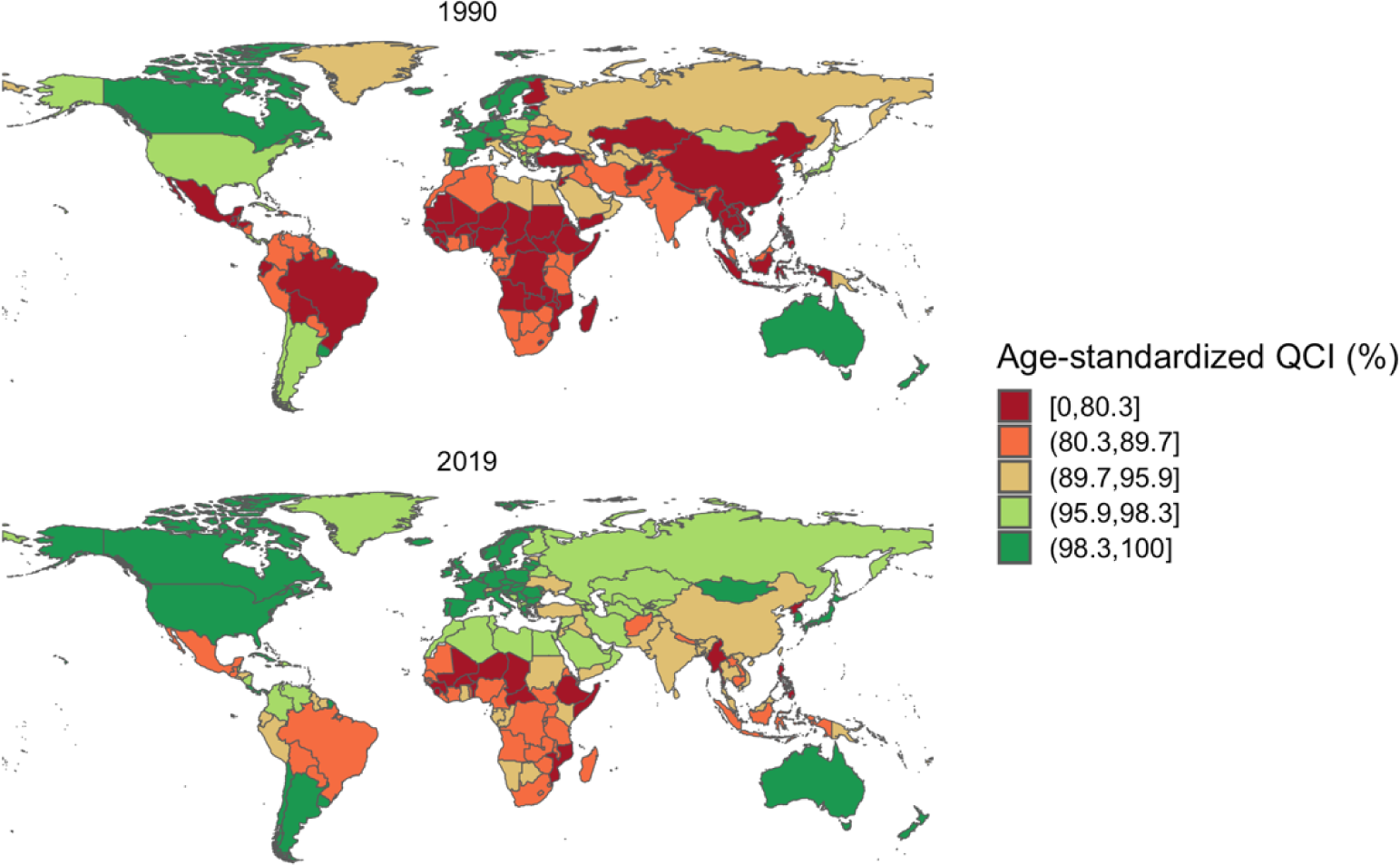
Geographical distribution of QCIs* (%) for lip and oral cavity cancer. A: Global distribution of age-standardized QCI in men and women in 1990, B: Global distribution of age-standardized QCI in men and women in 2019. QCI: Quality of care index.

**Table 1.** Estimates of burden and QCI of orofacial clefts by World Bank income groups.

| **DALYs rate in 2019**<br>**(per 100,000)** | **DALYs rate change**<br>**1990 to 2017** | **QCI in 2019 (%)** | **QCI change 1990 to**<br>**2019** |
| --- | --- | --- | --- |
| **World** |  |  |  |
| 7.51 (5.10–11.57) | –12.12 | 90.20 | 23.84 |
| **World Bank Regions** |  |  |  |
| High-income countries |  |  |  |
| 2.04 (1.29–2.95) | –1.00 | 98.68 | 3.25 |
| Upper-middle-income countries |  |  |  |
| 5.06 (3.88–6.57) | –25.10 | 92.88 | 54.55 |
| Lower-middle-income countries |  |  |  |
| 9.08 (6.16–13.45) | –8.18 | 91.17 | 11.70 |
| Low-income countries |  |  |  |
| 10.62 (5.29–22.14) | –5.12 | 80.33 | 11.26 |
| **SDI quintiles** |  |  |  |
| High SDI quintile |  |  |  |
| 2.20 (1.40–3.16) | –1.39 | 98.51 | 4.23 |
| High-middle SDI quintile |  |  |  |
| 4.66 (3.50–6.10) | –22.18 | 93.85 | 50.93 |
| Middle SDI quintile |  |  |  |
| 5.90 (4.33–7.80) | –14.55 | 93.04 | 29.67 |
| Low-middle SDI quintile |  |  |  |
| 9.30 (6.34–13.91) | –13.00 | 90.98 | 18.19 |
| Low SDI quintile |  |  |  |
| 10.97 (6.13–20.87) | –6.55 | 84.72 | 9.94 |

## Discussion

We applied the GBD data to a novel, multivariate index (the QCI) to assess the quality of care for orofacial clefts in both sexes and various regions worldwide from 1990 to 2019. Alteration patterns and trends were evaluated as well.

Several risk factors have been proposed for orofacial clefts: sex, race, ethnicity, environmental factors (e.g., maternal smoking and nutritional habits), and genetic factors (2, 20–24). Specific genetic variances (e.g., transforming growth factor-alpha polymorphisms) can substantially increase one’s susceptibility to the occurrence of orofacial clefts (25, 26). According to a national cohort study, cleft palate alone usually arises from different morphogenetic events than cleft lip with or without cleft palate. Additionally, a prognostic value has been proposed for cleft type to foreshadow the recurrence pattern and type in the patients’ first-degree relatives.

Regardless of the orofacial cleft type, because of difficulties in ascertaining the number of orofacial malformations where the pregnancy is spontaneously aborted, the true incidence is immeasurable for cleft palate alone and cleft lip with or without cleft palate. As a result, birth prevalence is the preferred measure to evaluate the frequency of orofacial clefts at birth (2). Consequently, we considered the prevalence in our index to reach accurate estimations for the QCI in different countries and regions. We used a more comprehensive approach than the previous indices (e.g., HAQI), considering our inclusion of the incidence, prevalence, YLL, and YLDs.

According to our results, from 1990 to 2019, the QCI increased in both males and females globally. This rise happened most markedly in the upper-middle-income (World Bank classification) and high-middle SDI countries in these 29 years. These findings can be expounded by considering the increasing awareness and embracing of preventive interventions as effective measures to counter the burden of orofacial clefts on the populations (27). Additionally, high- and upper-middle-income countries are most likely to have a pretty welcoming attitude toward the most cost-effective and sustainable interventions that can reduce the prevalence and afflictions of these malformations. Our findings support this speculation as the top five countries with the highest QCI scores in 2019 were all high SDI countries (Norway, Ireland, France, Germany, and the Netherlands).

On the other hand, all countries with the lowest QCI scores (Somalia, Niger, Burkina Faso, Ethiopia, and Mali) were located in Africa. The QCI of Somalia was 50% lower than the global value. This shows the significant importance of the existence of the minimum infrastructure and resources to improve the quality of care and implement preventive approaches to reduce the burden of orofacial diseases in each country.

Orofacial clefts not only reduce the quality of life but also impose high costs on the patients’ households (28, 29). Infant feeding, for instance, might be difficult and may lead to malnourishment and even death in environments without early surgical interventions (30, 31). Additionally, orofacial clefts are often accompanied by significant morbidity due to speech and hearing difficulty. These afflictions are associated with increased morbidity, mortality, and healthcare costs throughout life (32). Specialized dental treatment, speech therapy, and complex surgical treatments (occasionally with multiple repetitions) are often time-consuming and expensive. The use of telemedicine as a new and cost-effective management strategy has recently been proposed for remote diagnosis and recommendation by specialists (33–35). Analyzing the efficiency and optimizing the cost-effectiveness of such new treatment strategies primarily lies in the healthcare systems, and they should consider these factors in the policymaking process.

Orofacial clefts can also affect the oral health-related quality of life, functional well-being, and social-emotional status (e.g., self-esteem) in patients with the condition (especially in younger patients) (18, 36–38). These malformations can directly detriment the health status as well. According to a meta-analysis by Antonarakis et al. in 2014, children with cleft lip with or without cleft palate more frequently suffer from caries in deciduous and permanent dentitions compared with children without orofacial clefts (39). This can consequently exacerbate oral disorders in orofacial cleft patients, compassing a more significant proportion of the burden on them. Hearing loss has also been found more commonly in CLP and CPO patients (40, 41). These factors contribute to the fact that countries and regions with a scarcity of resources naturally experience a more significant burden of orofacial clefts and lower quality of care than more affluent countries (13, 42).

In countries with available treatments, other than care accessibility, affordability may still be an issue (even in developed countries) (13, 43). These social inequalities need to be immediately addressed as a priority by health care systems.

The quality of care showed a rising trend in the better part of the world. However, some high-income as well as Somalia and Zimbabwe, experienced a decrease in the QCI. Granted, all these countries had QCI values higher than the global average, but the downtrend in the quality of care from 1990 to 2019 warns of a possible relegation of the orofacial clefts. This might be due to the restricted age group with mortality due to these malformations. To prevent this issue from perpetuating and exacerbating, the healthcare systems in these regions should heed and manage orofacial clefts more proactively.

The QCI showed a converging pattern between males and females in the 1990-2019 period worldwide. Both genders experienced a continuous increase in the QCI. Based on the GDR results, African, South American, and Asian countries improved in terms of gender disparity and closing the gender gaps for orofacial clefts.

Effective management of the burden of orofacial clefts, including improving the quality of care, requires cost-efficient screening programs worldwide. The International Perinatal Database of Typical Orofacial Clefts (IPDTOC) has been a clear step forward in this regard on an international scale (44). Nevertheless, many countries, particularly in Africa and Asia, still do not have national registries to contribute, and inequalities still exist. Population-representative data are of utmost importance for future policymaking on orofacial clefts in all countries. The availability, adequacy and cost of necessary treatment are necessary to be addressed as well.

### Limitations

Despite its name, our index (QCI) cannot directly reflect the quality of care for orofacial clefts. Considering the limitations in providing global, regional, and national datasets for monitoring the status of these malformations, we tried to include the most relevant population-level data that could help infer and generalize the results to reach conclusions about the quality of care. For instance, higher ratios of YLLs to YLDs indicate higher mortality of the disease, meaning that higher figures represent a worse-off status regarding the survival of cancer patients. This can imply a lower quality of care in its specific country or region, albeit not being a direct representative of the quality of care. What is more, our estimations were made based on the GBD 2019 data on the orofacial clefts. Accordingly, one should be cautious when applying our results as they are valid merely on national and global scales (and not on an individual, per-patient basis). Finally, the QCI scores are relative values (not absolute values), and their use is limited to comparisons within the GBD 2019 database.

## Conclusion

The quality of care showed an uprising trend on a global scale from 1990 to 2019. Almost all countries experienced an increase in QCI values in both males and females. Upper-middle SDI and upper-to-middle-income regions enjoyed the highest rise in QCI, while a few countries had a decreasing trend in the same period. In order to halt this issue from exacerbating, these regions need to address the orofacial clefts more proactively in the future. Providing cost-effective preventive approaches (screening programs, altering nutrition behaviour, etc.) and the minimum infrastructure for the surgical treatment of orofacial clefts is indispensable. This is especially salient in African and LMIC countries.

## Data Availability

All code and data related to the study were shared via its OSF repository (https://osf.io/r94ch/) and GitHub (https://github.com/choxos/qci-cleft) at the time of submission of the manuscript.

## Notes

**Conflict of interest disclosure:** The authors declare no conflict of interest.

**Funding disclosure:**This study did not receive any funding.

### Competing Interest Statement

The authors have declared no competing interest.

### Clinical Protocols

https://osf.io/94wr7

### Funding Statement

This study did not receive any funding.

