## Appendix 1 for "Global, Regional, and National Survey on Burden and Quality of Care Index (QCI) of Orofacial Clefts: Global Burden of Disease Systematic Analysis 1990–2019"

### QCI of Orofacial Clefts - analysis

Ahmad Sofi-Mahmudi

2024-03-24

#### Loading required packages

```
pacman::p_load(dplyr,
               ggplot2,
               knitr,
               tidyr,
               readstata13,
               here,
               gt)
```

#### Loading the dataset

```
main_data = read.dta13(here("data", "main_data.dta"))
iso3 = read.dta13(here("data", "ISO3.dta"))
```

#### Results

Global Quality of Care (QCI):

```
kable(main_data %>%
        select(location_name, year, sex_name, pca_score) %>%
        filter(location_name == "Global",
                     year == c(1990, 2019)) %>%
        mutate(across(4, round, 1)))
```

```
## Warning: There was 1 warning in `mutate()`.
## i In argument: `across(4, round, 1)`.
## Caused by warning:
## ! The `...` argument of `across()` is deprecated as of dplyr 1.1.0.
## Supply arguments directly to `.fns` through an anonymous function instead.
##
##   # Previously
##   across(a:b, mean, na.rm = TRUE)
##
##   # Now
##   across(a:b, \(x) mean(x, na.rm = TRUE))
```

| location_name | year | sex_name | pca_score |
|---|---|---|---|
| Global | 1990 | Both | 66.4 |
| Global | 2019 | Both | 90.2 |

| location_name | year | sex_name | pca_score |
|---|---|---|---|
| Global | 1990 | Female | 82.9 |
| Global | 2019 | Female | 94.3 |
| Global | 1990 | Male | 72.8 |
| Global | 2019 | Male | 93.6 |

And its plot:

```
main_data %>%
        filter(location_name == "Global" & age_name == "Age-standardized") %>%
        ggplot(aes(year, pca_score, group = sex_name, color = sex_name)) +
        geom_line(size = 1, alpha = 0.8) +
        geom_point(size = 2) +
        scale_color_brewer(name = "Sex", palette = "Set1") +
        xlab("Year") +
        ylab("Quality of Care Index (QCI)")
```

```
## Warning: Using `size` aesthetic for lines was deprecated in ggplot2 3.4.0.
## i Please use `linewidth` instead.
## This warning is displayed once every 8 hours.
## Call `lifecycle::last_lifecycle_warnings()` to see where this warning was
## generated.
```

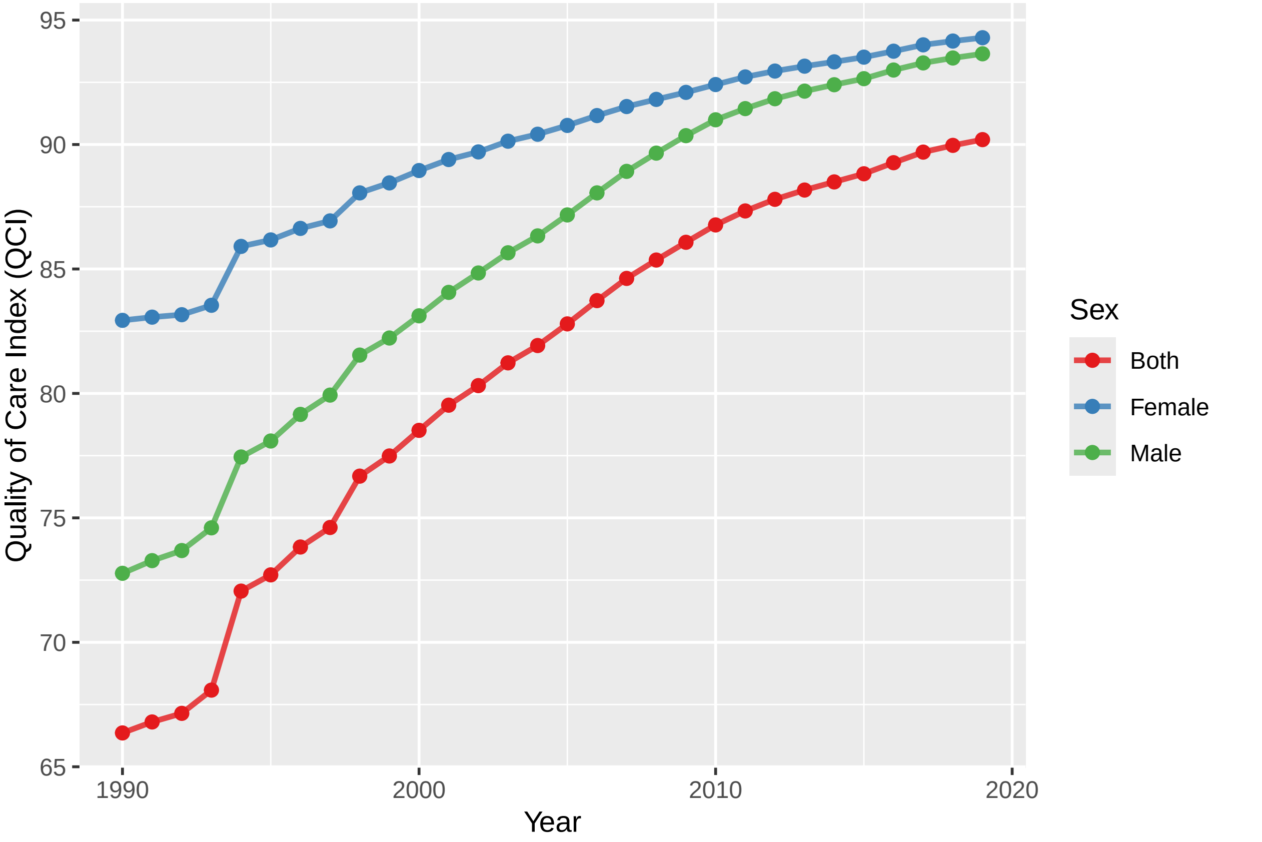


```
# dev.copy(tiff, "figures/Figure1.tiff", width = 20, height = 10, units = "cm", res = 300)
# dev.off()
```

Top countries:

```
kable(head(main_data %>%
        filter(year == 2019 & sex_name == "Both") %>%
        select(location_name, pca_score)  %>%
        arrange(desc(pca_score)) %>%
        mutate(across(2, round, 1))
     ))
```

| location_name | pca_score |
|---|---|
| Norway | 99.9 |
| Ireland | 99.4 |
| France | 99.4 |
| Germany | 99.3 |
| Netherlands | 99.3 |
| Malta | 99.3 |

Countries with the least QCI:

```
kable(tail(main_data %>%
        filter(year == 2019 & sex_name == "Both") %>%
        select(location_name, pca_score)  %>%
        arrange(desc(pca_score)) %>%
        mutate(across(2, round, 1))
     ))
```

| | location_name | pca_score |
|---|---|---|
| 217 | Sierra Leone | 75.3 |
| 218 | Mali | 74.4 |
| 219 | Ethiopia | 73.0 |
| 220 | Burkina Faso | 72.6 |
| 221 | Niger | 67.6 |
| 222 | Somalia | 59.1 |

Trend of change in QCI:

```
qci_trend = merge(main_data, iso3, by = "location_id") %>%
        select(location_name.y, year, sex_name, pca_score, type, iso3_countries) %>%
        filter(year %in% c(1990, 2019)) %>%
        distinct() %>%
        spread(year, pca_score) %>%
        mutate(year_ratio = `2019`/`1990`,
               compare = ifelse(`2019` > `1990`, "higher", "lower")) %>%
        mutate(year_ratio_cat = ifelse(year_ratio < 0.95, "lower",
                                       ifelse(year_ratio > 1.05, "higher", "equal")))
```

Countries in which the QCI has decreased:

```
kable(qci_trend %>%
        filter(sex_name == "Both" & type == "Country" & compare == "lower"))
```

| location_name.y | sex_name | type | iso3_countries | 1990 | 2019 | year_ratio | compare | year_ratio_cat |
|---|---|---|---|---|---|---|---|---|
| Albania | Both | Country | ALB | 98.07684 | 97.30689 | 0.9921495 | lower | equal |

| location_name.y | sex_name | type | iso3_countries | 1990 | 2019 | year_ratio | compare | year_ratio_cat |
|---|---|---|---|---|---|---|---|---|
| Guam | Both | Country | GUM | 96.41681 | 96.15187 | 0.9972522 | lower | equal |
| New Zealand | Both | Country | NZL | 99.06335 | 98.84734 | 0.9978196 | lower | equal |
| Northern Mariana Islands | Both | Country | MNP | 96.36154 | 96.27993 | 0.9991530 | lower | equal |
| Norway | Both | Country | NOR | 100.00000 | 99.90309 | 0.9990309 | lower | equal |
| Singapore | Both | Country | SGP | 99.23507 | 99.19500 | 0.9995961 | lower | equal |
| Somalia | Both | Country | SOM | 59.34079 | 59.05250 | 0.9951418 | lower | equal |
| Zimbabwe | Both | Country | ZWE | 85.88404 | 81.13136 | 0.9446616 | lower | lower |

```
qci_trend$type2 = factor(qci_trend$type, levels = c("Global", "WHO Regions", "World Bank Income Levels"

rows = (seq(nrow(filter(qci_trend, sex_name == "Both")))-1) %/% 37

qci_trend %>%
        dplyr::rename(Scale = type2, Location = location_name.y, QCI = `2019`) %>%
        filter(sex_name == "Both") %>%
        select(Scale, Location, QCI) %>%
        arrange(Scale, desc(QCI)) %>%
        mutate(QCI = round(QCI, 1)) %>%
        split(rows) %>%
        do.call(cbind, .) %>%
        #dplyr::rename(Scale = c(1, 4, 7, 10, 13), Location = c(2, 5, 8, 11, 14), QCI = c(3, 6, 9, 12, 
        gt() %>%
          cols_label(
                ends_with("QCI") ~ "QCI",
                ends_with("Scale") ~ "Scale",
                ends_with("Location") ~ "Location") %>%
        data_color(
                columns = c(`0.QCI`, `1.QCI`, `2.QCI`, `3.QCI`, `4.QCI`, `5.QCI`),
                method = "numeric",
                palette = c("red", "green"),
                domain = c(59.1, 99.9)) %>%
        data_color(
                columns = c(`0.Scale`, `1.Scale`, `2.Scale`, `3.Scale`, `4.Scale`, `5.Scale`),
                method = "factor",
                palette = "Set2"
        ) %>%
        gtsave("fig.png")


qci_trend %>%
        filter(sex_name == "Both") %>%
        select(location_name.y, type2, `2019`) %>%
        arrange(type2, desc(`2019`)) #%>%
```

```
##                       location_name.y                      type2     2019
## 1                              Global                     Global 90.19779
## 2                     European Region                WHO Regions 97.11111
## 3              Region of the Americas                WHO Regions 93.38609
## 4        Eastern Mediterranean Region                WHO Regions 92.02133
## 5              South-East Asia Region                WHO Regions 91.62646
## 6              Western Pacific Region                WHO Regions 91.29025
```

```
## 7                     African Region           WHO Regions 82.94872
## 8             World Bank High Income World Bank Income Levels 98.68135
## 9     World Bank Upper Middle Income World Bank Income Levels 92.88296
## 10    World Bank Lower Middle Income World Bank Income Levels 91.16962
## 11             World Bank Low Income World Bank Income Levels 80.33064
## 12                          High SDI                       SDI 98.51045
## 13                   High-middle SDI                       SDI 93.84630
## 14                        Middle SDI                       SDI 93.03736
## 15                    Low-middle SDI                       SDI 90.98462
## 16                           Low SDI                       SDI 84.72268
## 17                            Europe           4 World Regions 96.87435
## 18                           America           4 World Regions 93.38609
## 19                              Asia           4 World Regions 91.74093
## 20                            Africa           4 World Regions 84.87962
## 21                     North America        World Bank Regions 98.58294
## 22        Europe & Central Asia - WB        World Bank Regions 97.08795
## 23   Middle East & North Africa - WB        World Bank Regions 95.66187
## 24    Latin America & Caribbean - WB        World Bank Regions 92.37971
## 25                  South Asia - WB        World Bank Regions 91.82220
## 26          East Asia & Pacific - WB        World Bank Regions 90.30872
## 27           Sub-Saharan Africa - WB        World Bank Regions 82.38047
## 28                            Norway                   Country 99.90309
## 29                           Ireland                   Country 99.37874
## 30                            France                   Country 99.35528
## 31                           Germany                   Country 99.34695
## 32                       Netherlands                   Country 99.33693
## 33                             Malta                   Country 99.33319
## 34                             Spain                   Country 99.33005
## 35                          Portugal                   Country 99.32887
## 36                           Uruguay                   Country 99.31392
## 37                            Israel                   Country 99.29824
## 38                             Chile                   Country 99.28937
## 39                             Italy                   Country 99.27780
## 40                            Canada                   Country 99.26611
## 41                           Denmark                   Country 99.26362
## 42                           Austria                   Country 99.26275
## 43                           Iceland                   Country 99.25965
## 44                        Luxembourg                   Country 99.25373
## 45                         Australia                   Country 99.25336
## 46                           Belgium                   Country 99.24154
## 47                           Andorra                   Country 99.20224
## 48                            Sweden                   Country 99.19927
## 49                         Singapore                   Country 99.19500
## 50                    United Kingdom                   Country 99.15592
## 51                            Greece                   Country 99.15589
## 52                          Slovenia                   Country 99.12773
## 53                           Hungary                   Country 99.09388
## 54                         Argentina                   Country 99.08915
## 55                    Czech Republic                   Country 99.05188
## 56                           Croatia                   Country 99.03882
## 57                          Bulgaria                   Country 99.02584
## 58                         Lithuania                   Country 98.98025
## 59                            Brunei                   Country 98.97715
## 60                            Latvia                   Country 98.96509
```

```
## 61                      South Korea                  Country 98.92488
## 62                           Panama                  Country 98.89643
## 63                          Romania                  Country 98.87222
## 64                      New Zealand                  Country 98.84734
## 65                       Costa Rica                  Country 98.83433
## 66                          Moldova                  Country 98.82697
## 67                           Cyprus                  Country 98.82615
## 68                            Japan                  Country 98.69619
## 69                           Poland                  Country 98.65582
## 70                           Serbia                  Country 98.62487
## 71                    United States                  Country 98.50286
## 72                      Puerto Rico                  Country 98.41362
## 73                          Bermuda                  Country 98.39093
## 74                       Montenegro                  Country 98.35171
## 75                         Slovakia                  Country 98.34010
## 76                      The Bahamas                  Country 98.33795
## 77              Trinidad and Tobago                  Country 98.32139
## 78                             Cuba                  Country 98.31545
## 79                         Mongolia                  Country 98.30684
## 80                       Uzbekistan                  Country 98.29204
## 81           Bosnia and Herzegovina                  Country 98.25654
## 82                          Belarus                  Country 98.19649
## 83                          Finland                  Country 98.14070
## 84                       Kazakhstan                  Country 98.09598
## 85              Virgin Islands, U.S.                 Country 98.09018
## 86                       Azerbaijan                  Country 98.08890
## 87                          Georgia                  Country 98.00003
## 88                          Grenada                  Country 97.98003
## 89                         Colombia                  Country 97.95313
## 90                       Kyrgyzstan                  Country 97.92165
## 91                        Macedonia                  Country 97.90547
## 92                      Saint Lucia                  Country 97.90209
## 93                         Barbados                  Country 97.88090
## 94                        Mauritius                  Country 97.76093
## 95                        Greenland                  Country 97.75769
## 96                     Turkmenistan                  Country 97.64197
## 97                           Kuwait                  Country 97.63782
## 98                          Jamaica                  Country 97.57385
## 99               Dominican Republic                  Country 97.41686
## 100                         Albania                  Country 97.30689
## 101                           Qatar                  Country 97.26739
## 102            United Arab Emirates                  Country 97.13076
## 103                            Oman                  Country 97.03222
## 104              Russian Federation                  Country 97.03115
## 105                    Saudi Arabia                  Country 96.96500
## 106                         Armenia                  Country 96.88999
## 107                       Venezuela                  Country 96.87538
## 108                           Syria                  Country 96.64943
## 109                         Bahrain                  Country 96.63952
## 110                  American Samoa                  Country 96.58036
## 111                          Belize                  Country 96.56692
## 112                           Egypt                  Country 96.51796
## 113                         Lebanon                  Country 96.46144
## 114                       Palestine                  Country 96.37350
```

```
## 115                          Morocco                  Country 96.32884
## 116         Northern Mariana Islands                  Country 96.27993
## 117                            Libya                  Country 96.21758
## 118                             Guam                  Country 96.15187
## 119                            Tonga                  Country 96.14428
## 120                       Tajikistan                  Country 96.05603
## 121                          Algeria                  Country 96.03327
## 122   Federated States of Micronesia                  Country 95.92559
## 123                          Tunisia                  Country 95.91315
## 124                             Iran                  Country 95.89016
## 125                        Nicaragua                  Country 95.85928
## 126                             Iraq                  Country 95.85741
## 127                 Marshall Islands                  Country 95.84155
## 128                             Fiji                  Country 95.76432
## 129                      El Salvador                  Country 95.76380
## 130                            Samoa                  Country 95.76228
## 131 Saint Vincent and the Grenadines                  Country 95.71629
## 132                          Vanuatu                  Country 95.69464
## 133                          Estonia                  Country 95.62623
## 134                  Solomon Islands                  Country 95.61161
## 135                         Suriname                  Country 95.34810
## 136                             Peru                  Country 95.28146
## 137                           Guyana                  Country 95.27401
## 138                Equatorial Guinea                  Country 95.17824
## 139                         Kiribati                  Country 95.00594
## 140                 Papua New Guinea                  Country 94.89874
## 141                          Ecuador                  Country 94.87318
## 142                            Gabon                  Country 94.63134
## 143                          Ukraine                  Country 94.48456
## 144              Antigua and Barbuda                  Country 94.00753
## 145                        Sri Lanka                  Country 93.54921
## 146                            Sudan                  Country 93.53246
## 147                           Taiwan                  Country 93.39876
## 148                           Bhutan                  Country 92.85331
## 149                      Switzerland                  Country 92.63828
## 150                         Botswana                  Country 92.41345
## 151                         Honduras                  Country 92.41233
## 152                         Thailand                  Country 92.36613
## 153                            India                  Country 92.31184
## 154                         Dominica                  Country 92.11439
## 155                           Turkey                  Country 92.04578
## 156                          Vietnam                  Country 91.79236
## 157                            Congo                  Country 91.59420
## 158                           Jordan                  Country 91.55416
## 159                            Yemen                  Country 91.53292
## 160                            China                  Country 91.51940
## 161                       Seychelles                  Country 91.48093
## 162                         Pakistan                  Country 91.45064
## 163                       Cape Verde                  Country 91.35201
## 164                       Bangladesh                  Country 91.19101
## 165                          Namibia                  Country 90.95483
## 166                            Ghana                  Country 90.92936
## 167                            Haiti                  Country 90.49722
## 168                         Maldives                  Country 90.35650
```

```
## 169                         Malaysia                  Country 90.21110
## 170                            Kenya                  Country 89.84286
## 171            Sao Tome and Principe                  Country 89.72025
## 172                           Mexico                  Country 89.66448
## 173                            Nepal                  Country 89.61565
## 174                           Uganda                  Country 89.43847
## 175                       Mauritania                  Country 89.36026
## 176                         Cameroon                  Country 89.31353
## 177                           Angola                  Country 89.16236
## 178                    Cote d'Ivoire                  Country 89.07287
## 179                        Guatemala                  Country 88.96181
## 180                           Rwanda                  Country 88.56727
## 181                        Swaziland                  Country 88.32760
## 182                           Zambia                  Country 87.59568
## 183                          Bolivia                  Country 87.29250
## 184                     South Africa                  Country 87.23564
## 185                           Brazil                  Country 87.10227
## 186                             Togo                  Country 86.92593
## 187                      Afghanistan                  Country 86.20671
## 188                          Comoros                  Country 86.12101
## 189                         Djibouti                  Country 85.88850
## 190                       Madagascar                  Country 85.81094
## 191                          Liberia                  Country 85.58770
## 192                           Malawi                  Country 85.32916
## 193                        Indonesia                  Country 85.21461
## 194                         Tanzania                  Country 84.89089
## 195                          Lesotho                  Country 84.80394
## 196                    Guinea-Bissau                  Country 84.45294
## 197 Democratic Republic of the Congo                  Country 83.65864
## 198                       The Gambia                  Country 83.60179
## 199                          Nigeria                  Country 82.95918
## 200                          Eritrea                  Country 82.83619
## 201                         Paraguay                  Country 82.71399
## 202                          Burundi                  Country 82.69759
## 203                      Timor-Leste                  Country 82.25290
## 204                          Senegal                  Country 82.11638
## 205                         Cambodia                  Country 81.64107
## 206                         Zimbabwe                  Country 81.13136
## 207                      South Sudan                  Country 80.80632
## 208                             Laos                  Country 80.48815
## 209                      Philippines                  Country 79.89827
## 210                            Benin                  Country 79.76445
## 211                          Myanmar                  Country 79.70363
## 212         Central African Republic                  Country 77.02683
## 213                      North Korea                  Country 76.79888
## 214                           Guinea                  Country 76.79844
## 215                       Mozambique                  Country 75.70680
## 216                             Chad                  Country 75.49664
## 217                     Sierra Leone                  Country 75.27606
## 218                             Mali                  Country 74.44304
## 219                         Ethiopia                  Country 73.00839
## 220                     Burkina Faso                  Country 72.59247
## 221                            Niger                  Country 67.62856
## 222                          Somalia                  Country 59.05250
```

```r
#         group_map(~ qci_table(.x)) %>%
#         data.frame(.) %>%
#         gt() %>%
#         fmt_markdown(columns = TRUE)
```

Computing gender disparity ratio (GDR):

```r
gdr = merge(main_data, iso3, by = "location_name") %>%
        select(location_name, year, sex_name, pca_score, type, iso3_countries) %>%
        filter(sex_name %in% c("Male", "Female")) %>%
        distinct() %>%
        spread(sex_name, pca_score) %>%
        mutate(gdr = Female/Male) %>%
        mutate(gdr_cat = ifelse(gdr < 0.95, "low",
                                ifelse(gdr > 1.05, "high", "equal"))
               )
```

Comparing GDR categories in 1990 and 2019:

```r
kable(merge(gdr %>%
              filter(year == 1990 & type == "Country") %>%
              group_by(gdr_cat) %>%
              summarise(n = n()),
      gdr %>%
              filter(year == 2019 & type == "Country") %>%
              group_by(gdr_cat) %>%
              summarise(n = n()),
      by = "gdr_cat") %>% setNames(c("gdr_cat", "1990", "2019")))
```

| gdr_cat | 1990 | 2019 |
|---|---|---|
| equal | 123 | 163 |
| high | 29 | 9 |
| low | 21 | 1 |

Comparing SDI world regions in terms of equal GDR:

```r
kable(merge(gdr %>%
              filter(year == 1990 & type == "SDI") %>%
              group_by(location_name) %>%
              summarise(equal = ifelse(gdr_cat == "equal", TRUE, FALSE)),
      gdr %>%
              filter(year == 2019 & type == "SDI") %>%
              group_by(location_name) %>%
              summarise(equal = ifelse(gdr_cat == "equal", TRUE, FALSE)),
      by = "location_name") %>% setNames(c("location_name", "1990", "2019")))
```

| location_name | 1990 | 2019 |
|---|---|---|
| High SDI | TRUE | TRUE |
| High-middle SDI | FALSE | TRUE |
| Low SDI | TRUE | TRUE |
| Low-middle SDI | FALSE | TRUE |
| Middle SDI | FALSE | TRUE |

Comparing World Bank Income Levels in terms of equal GDR:

```
kable(merge(gdr %>%
              filter(year == 1990 & type == "World Bank Income Levels") %>%
              group_by(location_name) %>%
              summarise(equal = ifelse(gdr_cat == "equal", TRUE, FALSE)),
      gdr %>%
              filter(year == 2019 & type == "World Bank Income Levels") %>%
              group_by(location_name) %>%
              summarise(equal = ifelse(gdr_cat == "equal", TRUE, FALSE)),
      by = "location_name") %>% setNames(c("location_name", "1990", "2019")))
```

| location_name | 1990 | 2019 |
|---|---|---|
| World Bank High Income | TRUE | TRUE |
| World Bank Low Income | TRUE | TRUE |
| World Bank Lower Middle Income | FALSE | TRUE |
| World Bank Upper Middle Income | FALSE | TRUE |

### Loading required packages

```
pacman::p_load(clipr,
               tidyr,
               dplyr,
               readstata13,
               data.table,
               FactoMineR,
               foreign,
               RColorBrewer,
               rgdal,
               ggplot2,
               readxl,
               randomcoloR,
               viridis,
               ineq,
               sp,
               LSD,
               reshape,
               ff,
               ffbase)
```

```
## Warning: package 'rgdal' is not available for this version of R
##
## A version of this package for your version of R might be available elsewhere,
## see the ideas at
## https://cran.r-project.org/doc/manuals/r-patched/R-admin.html#Installing-packages

## Warning in p_install(package, character.only = TRUE, ...):

## Warning in library(package, lib.loc = lib.loc, character.only = TRUE,
## logical.return = TRUE, : there is no package called 'rgdal'

## Warning: package 'ffbase' is not available for this version of R
##
## A version of this package for your version of R might be available elsewhere,
```

```
## see the ideas at
## https://cran.r-project.org/doc/manuals/r-patched/R-admin.html#Installing-packages

## Warning in p_install(package, character.only = TRUE, ...):

## Warning in library(package, lib.loc = lib.loc, character.only = TRUE,
## logical.return = TRUE, : there is no package called 'ffbase'

## Warning in pacman::p_load(clipr, tidyr, dplyr, readstata13, data.table, : Failed to install/load:
## rgdal, ffbase
```

The datasets are now created. Next, we w Map

```r
#install.packages(c("cowplot", "googleway", "ggplot2", "ggrepel",
#                   "ggspatial", "libwgeom", "sf", "rnaturalearth", #"rnaturalearthdata", "rgeos", "hav

library("haven")
library("readstata13")
library("dplyr")
library("tidyr")
library("ggplot2")
theme_set(theme_bw())
library("sf")
```

```
## Linking to GEOS 3.11.0, GDAL 3.5.3, PROJ 9.1.0; sf_use_s2() is TRUE
```

```r
library("rnaturalearth")
library("rnaturalearthdata")
```

```
##
## Attaching package: 'rnaturalearthdata'

## The following object is masked from 'package:rnaturalearth':
##
##     countries110
```

```r
library("rgeos")
```

```
## rgeos version: 0.6-4, (SVN revision 699)
##  GEOS runtime version: 3.12.1-CAPI-1.18.1
##  Please note that rgeos will be retired during October 2023,
## plan transition to sf or terra functions using GEOS at your earliest convenience.
## See https://r-spatial.org/r/2023/05/15/evolution4.html for details.
##  GEOS using OverlayNG
##  Linking to sp version: 2.1-3
##  Polygon checking: TRUE

##
## Attaching package: 'rgeos'

## The following object is masked from 'package:bit':
##
##     symdiff

## The following object is masked from 'package:dplyr':
##
##     symdiff
```

```r
map_data = merge(main_data, iso3, by = 'location_id')
map_data$Location = map_data$location_name
map_data$QCI = map_data$pca
```

```
map_data$iso = map_data$iso3_countries
data2 = filter(map_data, year %in% c(1990, 2019), sex_name == 'Both', age_name == "Age-standardized", ty


quantile_values = quantile(data2$QCI, probs = seq(0, 1, 1/5))
data2$quantile = cut(data2$QCI, breaks = quantile_values, include.lowest = TRUE)

world = ne_countries(scale = "medium", returnclass = "sf")
world$iso = world$iso_a3
world = merge(world, data2, by = 'iso')


ggplot(data = world) +
        geom_sf(aes(fill = quantile)) +
        scale_fill_manual(values = c("#a40227", "#f56c42", "#dfbf72", "#a7da68", "#1a9850"), name = "Ag
        theme_void() +
        facet_wrap(~ year, ncol = 1) +
        theme(legend.key.size = unit(0.4, 'cm'))
```

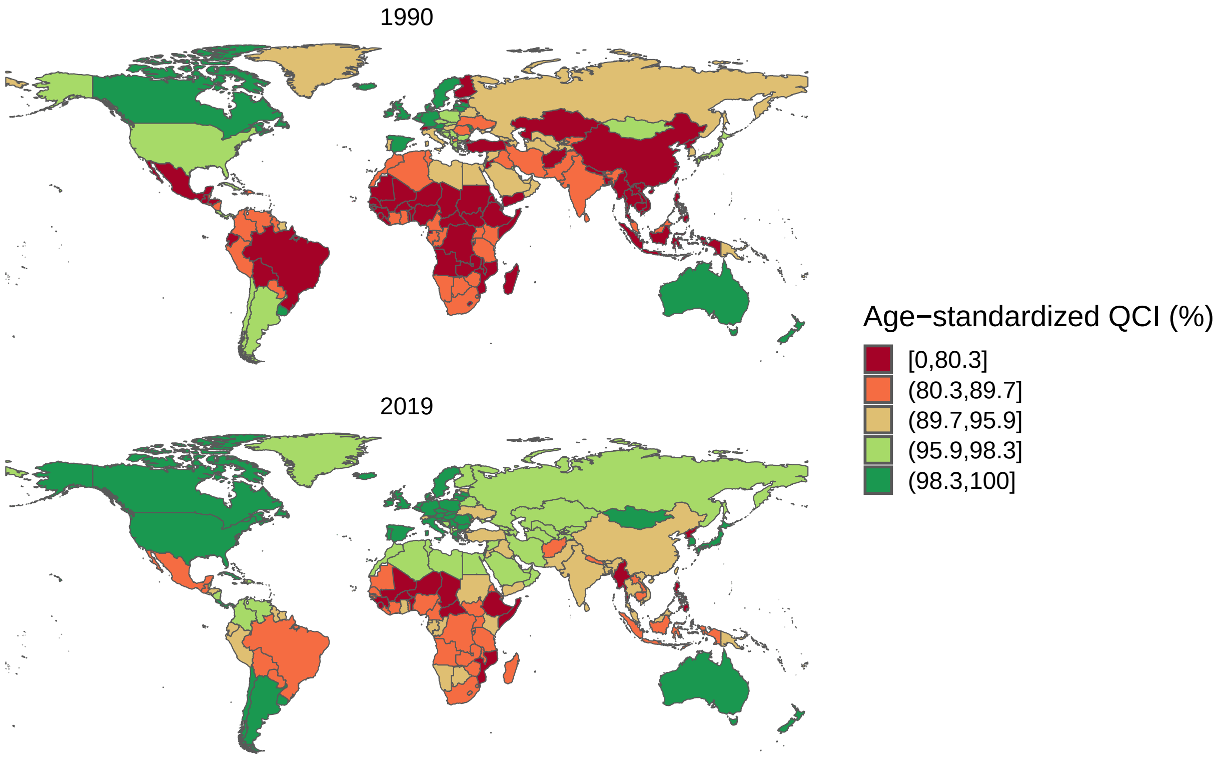


```
# dev.copy(tiff, "figures/Figure3.tiff", width = 20, height = 10, units = "cm", res = 300)
# dev.off()
```

GDR

```
# install.packages(c("cowplot", "googleway", "ggplot2", "ggrepel",
#                    "ggspatial", "libwgeom", "sf", "rnaturalearth", "rnaturalearthdata", "rgeos", "have

library("haven")
library("readstata13")
library("dplyr")
```

```r
library("tidyr")
library("ggplot2")
theme_set(theme_bw())
library("sf")
library("rnaturalearth")
library("rnaturalearthdata")
library("rgeos")
gdr_data = merge(main_data, iso3, by = 'location_id')
gdr_data$Location = gdr_data$location_id
gdr_data$QCI = gdr_data$pca
gdr_data$iso = gdr_data$iso3_countries
data2 = filter(gdr_data, year %in% c(1990, 2019), sex_name %in% c('Male', 'Female'), age_name == "Age-s
data3 = select(data2, location_id, iso, sex_name, year, QCI)
data4 = spread(data3, sex_name, QCI)
data4$GDR = data4$Female/data4$Male

breaks <- c(0, 0.5, 0.95, 1.05, 2.0, Inf)
labels <- c("< 0.5", "[0.5, 0.95)", "[0.95, 1.05]", "(1.05, 2.0]", "> 2.0")
data4$quantile <- cut(data4$GDR, breaks = breaks, labels = labels, include.lowest = TRUE)

world = ne_countries(scale = "medium", returnclass = "sf")
world$iso = world$iso_a3
world = merge(world, data4, by = 'iso')

ggplot(data = world) +
        geom_sf(aes(fill = quantile)) +
        scale_fill_manual(values = c("#878787", "#b7b7b7", "#64c2a5", "#b1abd0", "#7f74ac"), name = "Ag
        theme_void() +
        facet_wrap(~ year, ncol = 1) +
        theme(legend.key.size = unit(0.4, 'cm'))
```

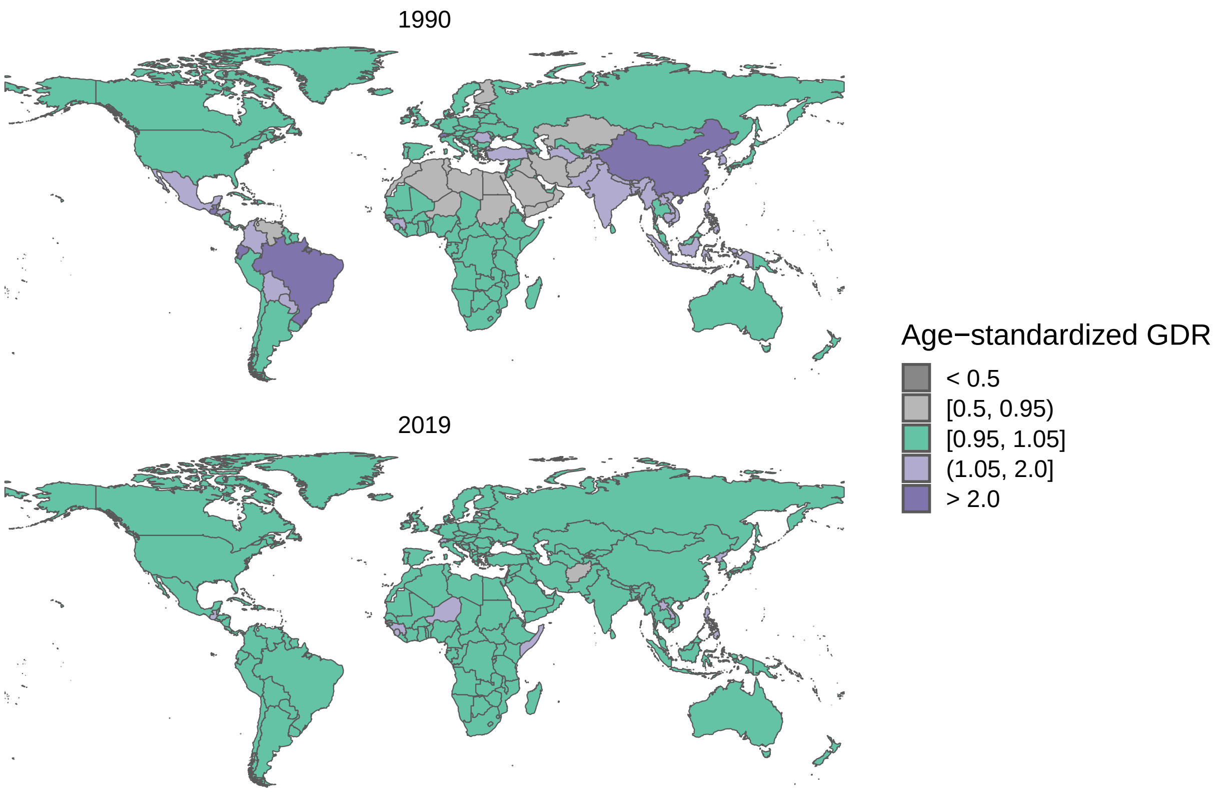


```r
# dev.copy(tiff, "figures/Figure2.tiff", width = 20, height = 10, units = "cm", res = 300)

dev.off()
```

```
## null device
##           1
```

```r
data3 = select(data2, Location, QCI)
```
